# Methotrexate Hampers Immunogenicity to BNT162b2 mRNA COVID-19 Vaccine in Immune-Mediated Inflammatory Disease

**DOI:** 10.1101/2021.05.11.21256917

**Authors:** Rebecca H. Haberman, Ramin Sedaghat Herati, David Simon, Marie Samanovic, Rebecca B. Blank, Michael Tuen, Sergei B. Koralov, Raja Atreya, Koray Tascilar, Joseph R. Allen, Rochelle Castillo, Amber R. Cornelius, Paula Rackoff, Gary Solomon, Samrachana Adhikari, Natalie Azar, Pamela Rosenthal, Peter Izmirly, Jonathan Samuels, Brian Golden, Soumya Reddy, Markus Neurath, Steven B. Abramson, Georg Schett, Mark J. Mulligan, Jose U. Scher

## Abstract

**Objective:** To investigate the humoral and cellular immune response to mRNA COVID-19 vaccines in patients with immune-mediated inflammatory diseases (IMIDs) on immunomodulatory treatment.

**Methods:** Established patients at NYU Langone Health with IMID (n=51) receiving the BNT162b2 mRNA vaccination were assessed at baseline and after second immunization. Healthy subjects served as controls (n=26). IgG antibody responses to the spike protein were analyzed for humoral response. Cellular immune response to SARS-CoV-2 was further analyzed using high-parameter spectral flow cytometry. A second independent, validation cohort of controls (n=182) and patients with IMID (n=31) from Erlangen, Germany were also analyzed for humoral immune response.

**Results:** Although healthy subjects (n=208) and IMID patients on biologic treatments (mostly on TNF blockers, n=37) demonstrate robust antibody responses (over 90%), those patients with IMID on background methotrexate (n=45) achieve an adequate response in only 62.2% of cases. Similarly, IMID patients do not demonstrate an increase in CD8+ T cell activation after vaccination.

**Conclusions:** In two independent cohorts of IMID patients, methotrexate, a widely used immunomodulator for the treatment of several IMIDs, adversely affected humoral and cellular immune response to COVID-19 mRNA vaccines. Although precise cut offs for immunogenicity that correlate with vaccine efficacy are yet to be established, our findings suggest that different strategies may need to be explored in patients with IMID taking methotrexate to increase the chances of immunization efficacy against SARS-CoV-2 as has been demonstrated for augmenting immunogenicity to other viral vaccines.

**KEY MESSAGES:** *What is already known about this subject?:*

- The impact of COVID-19 has been felt across the globe and new hope has arisen with the approval of mRNA vaccines against the SARS-CoV-2. Studies have shown immunogenicity and efficacy rates of over 90% in the immunocompetent adult population. However, there is a lack of knowledge surrounding the response of patients with immune-mediated inflammatory diseases (IMIDs) who may also be on immunomodulatory medications.
- Patients with IMID have been shown to have attenuated immune responses to seasonal influenza vaccination.

*What does this study add?:*

- This study looks at the humoral and cellular immune response to two doses of BNT162b2 mRNA COVID-19 Vaccine in participants with IMID (on immunomodulators) compared with healthy controls.
- Individuals with IMID on methotrexate demonstrate up to a 62% reduced rate of adequate immunogenicity to the BNT162b2 mRNA vaccination. Those on anti-cytokine or non-methotrexate oral medications demonstrate similar levels of immunogenicity as healthy controls (greater than 90%).
- Similarly, vaccination did not induce an activated CD8+ T cell response in participants on background methotrexate, unlike healthy controls and patients with IMID not receiving methotrexate.

*How might this impact of clinical practice or future developments?:*

- These results suggest that patients on methotrexate may need alternate vaccination strategies such as additional doses of vaccine, dose modification of methotrexate, or even a temporary discontinuation of this drug. Further studies will be required to explore the effect of these approaches on mRNA vaccine immunogenicity.

## INTRODUCTION

Patients with immune-mediated inflammatory diseases (IMIDs), have an inherently heightened susceptibility to infection and may thus be considered high risk for developing *coronavirus disease 2019 (COVID-19)*. Importantly, however, the strength of response to viral vaccines (i.e., influenza and hepatitis B) and their long-lasting protective effects in IMID patients taking conventional-disease modifying anti-rheumatic (cDMARDs), such as methotrexate, or biologic (bDMARDs), such as tumor necrosis factor inhibitors (TNFis), may not be as robust as it is in the general population following immunization.(1-5) Data regarding mRNA COVID-19 vaccines’ safety, immunogenicity and efficacy are rapidly emerging for the immunocompetent adult population,(6) where more than 90% subjects achieve a satisfactory humoral response. However, the ability of IMID patients to adequately respond to these vaccines and the differences in humoral and cellular immune response to SARS-CoV-2 vaccination are not known, leaving a significant gap in knowledge that prevents optimal management of this patient population.

Given the experience with seasonal influenza vaccine immunogenicity,(2, 7) we hypothesized that IMID patients treated chronically with certain csDMARDs (i.e., methotrexate) would have an attenuated response to mRNA COVID-19 vaccines compared to IMID patients receiving anti-cytokine treatment or to non-IMID participants. To achieve this, we obtained pre- and post-immunization peripheral blood monocyte cells (PBMCs) and sera from IMID participants (n=82) in two independent cohorts (SAGA (Serologic Testing And Genomic Analysis of Autoimmune, Immune-Mediated, and Rheumatic Patients with COVID-19) cohort in New York City, USA and Erlangen cohort, Germany) and analyzed SARS-CoV-2-spike-specific antibody titers as compared to non-IMID controls (n=208). Cellular immune responses were further investigated using high-dimensional spectral flow cytometry in the New York City cohort.

## METHODS

### Participants

Established patients with IMID (n=51) receiving methotrexate, anti-cytokine biologics, or both participating in the SAGA study at New York University Langone Health in New York City,(8) who were receiving BNT162b2 mRNA vaccination were assessed at baseline and after the second dose during the period from December 23, 2020 through March 31, 2021. Healthy subjects served as controls (n= 26). IgG antibody responses to the S protein were analyzed for humoral immune response. A second independent validation cohort of controls (n=182) and patients with IMID (n=31) on either TNFi or methotrexate monotherapy from Erlangen, Germany was also analyzed for humoral response. Cellular immune responses to the vaccine were also studied for the New York SAGA participants using high-parameter spectral flow cytometry.

### Humoral and cellular immune response to BNT162b2 mRNA Vaccine

Humoral immune response was assessed by testing IgG antibodies against the spike protein of SAR-CoV-2.(9) In the New York Cohort, direct ELISA was used to quantify antibody titers on serum as previously described.(10) Titer of 5,000 units or greater was used as the cut off to determine an adequate response to vaccination. IgG antibodies against the S1 domain of the spike protein of SARS-CoV-2 were tested in the Erlangen participants using the commercial enzyme–linked immunosorbent assay from Euroimmun (Lübeck, Germany) on the EUROIMMUN Analyzer I platform and according to the manufacturers protocol.(11) Adequate response was defined as greater than 5.7 nm OD. Immune cell phenotyping pre- and post-immunization in New York participants was performed by 35-color spectral flow cytometry on PBMCs. Further details on methodology and analysis can be found in the Supplementary Appendix.

### Statistical Analysis

Patient characteristics were summarized using means, medians, standard deviations, ranges, and percentages as appropriate. Chi squared tests of independence and Fischer’s exact tests were used for categorical data. Mann Whitney U and Kruskal Wallis tests were used for unpaired continuous data and Wilcoxon Rank Sum tests were used for paired continuous data. A *p* value of less than .05 was considered significant. All analyses were done using R v3.6.0 software (R Foundation for Statistical Computing) and GraphPad Prism v9 (GraphPad Software).

### Patient and Public Involvement

This study was designed in response to frequent questions asked by patients with IMID but did not contain any direct public involvement.

## RESULTS

The New York cohort was comprised of 26 healthy individuals, 25 individuals with IMID receiving methotrexate monotherapy or in combination with other immunomodulatory medications, and 26 individuals with IMID on anti-cytokine therapy and/or other oral immunomodulators (Table 1). Healthy individuals and those with IMID not on methotrexate were similar in age (49.2 +/- 11.9 and 49.1 +/- 14.9 years respectively), while those IMID patients receiving methotrexate were generally older (63.2 +/- 11.9). IMID diagnoses were predominantly psoriasis/psoriatic arthritis and rheumatoid arthritis. The Erlangen cohort consisted of 182 healthy subjects, 11 subjects with IMID receiving TNFi monotherapy, and 20 subjects with IMID on methotrexate monotherapy (Supplementary Table 1). Individuals on methotrexate monotherapy were on average older than healthy individuals and those with IMID not on methotrexate (54.5 +/- 19.2 vs. 40.8+/- 12 .0 and 45.0 +/- 15.5 respectively).

**Table 1.**
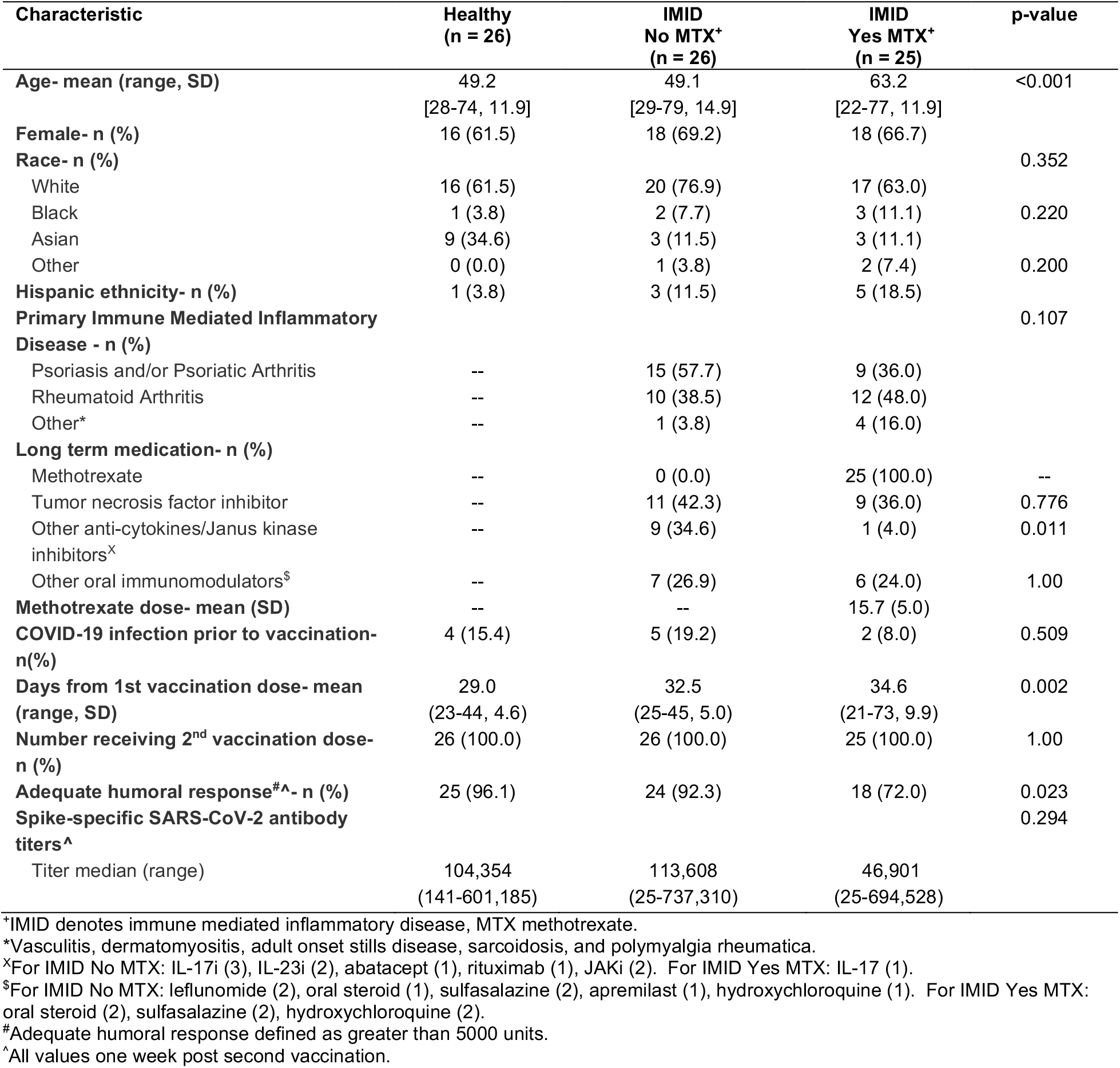
Baseline characteristics and spike-specific SARS-CoV-2 antibody titers in the New York Cohort.

| Characteristic | Healthy<br>(n = 26) | IMID<br>No MTX<sup>+</sup><br>(n = 26) | IMID<br>Yes MTX<sup>+</sup><br>(n = 25) | p-value |
| --- | --- | --- | --- | --- |
| **Age- mean (range, SD)** | 49.2<br>[28-74, 11.9] | 49.1<br>[29-79, 14.9] | 63.2<br>[22-77, 11.9] | <0.001 |
| **Female- n (%)** | 16 (61.5) | 18 (69.2) | 18 (66.7) |  |
| **Race- n (%)** |  |  |  | 0.352 |
| White | 16 (61.5) | 20 (76.9) | 17 (63.0) |  |
| Black | 1 (3.8) | 2 (7.7) | 3 (11.1) | 0.220 |
| Asian | 9 (34.6) | 3 (11.5) | 3 (11.1) |  |
| Other | 0 (0.0) | 1 (3.8) | 2 (7.4) | 0.200 |
| **Hispanic ethnicity- n (%)** | 1 (3.8) | 3 (11.5) | 5 (18.5) |  |
| **Primary Immune Mediated Inflammatory Disease - n (%)** |  |  |  | 0.107 |
| Psoriasis and/or Psoriatic Arthritis | -- | 15 (57.7) | 9 (36.0) |  |
| Rheumatoid Arthritis | -- | 10 (38.5) | 12 (48.0) |  |
| Other* | -- | 1 (3.8) | 4 (16.0) |  |
| **Long term medication- n (%)** |  |  |  |  |
| Methotrexate | -- | 0 (0.0) | 25 (100.0) | -- |
| Tumor necrosis factor inhibitor | -- | 11 (42.3) | 9 (36.0) | 0.776 |
| Other anti-cytokines/Janus kinase inhibitors<sup>x</sup> | -- | 9 (34.6) | 1 (4.0) | 0.011 |
| Other oral immunomodulators<sup>s</sup> | -- | 7 (26.9) | 6 (24.0) | 1.00 |
| **Methotrexate dose- mean (SD)** | -- | -- | 15.7 (5.0) |  |
| **COVID-19 infection prior to vaccination- n(%)** | 4 (15.4) | 5 (19.2) | 2 (8.0) | 0.509 |
| **Days from 1st vaccination dose- mean (range, SD)** | 29.0<br>(23-44, 4.6) | 32.5<br>(25-45, 5.0) | 34.6<br>(21-73, 9.9) | 0.002 |
| **Number receiving 2<sup>nd</sup> vaccination dose- n (%)** | 26 (100.0) | 26 (100.0) | 25 (100.0) | 1.00 |
| **Adequate humoral response<sup>#</sup>- n (%)** | 25 (96.1) | 24 (92.3) | 18 (72.0) | 0.023 |
| **Spike-specific SARS-CoV-2 antibody titers<sup>^</sup>** |  |  |  | 0.294 |
| Titer median (range) | 104,354<br>(141-601,185) | 113,608<br>(25-737,310) | 46,901<br>(25-694,528) |  |

\*IMID denotes immune mediated inflammatory disease, MTX methotrexate.
\*Vasculitis, dermatomyositis, adult onset stills disease, sarcoidosis, and polymyalgia rheumatica.
<sup>x</sup>For IMID No MTX: IL-17i (3), IL-23i (2), abatacept (1), rituximab (1), JAKi (2). For IMID Yes MTX: IL-17 (1).
<sup>s</sup>For IMID No MTX: leflunomide (2), oral steroid (1), sulfasalazine (2), apremilast (1), hydroxychloroquine (1). For IMID Yes MTX: oral steroid (2), sulfasalazine (2), hydroxychloroquine (2).
<sup>#</sup>Adequate humoral response defined as greater than 5000 units.
<sup>^</sup>All values one week post second vaccination.

### Decreased antibody response to mRNA COVID-19 vaccine in IMID patients on methotrexate

Immunogenicity was characterized by testing IgG antibodies against the spike protein of SARS-CoV-2. In the New York cohort, of the healthy participants, 25 of 26 (96.1%) demonstrated adequate humoral immune response. Patients with IMID not on methotrexate achieved a similar rate of high antibody titers (24/26, 92.3%), while those on methotrexate had a lower rate of adequate humoral response (18/25, 72.0%) (Figure 1A, Table 1). This remains true even after the exclusion of patients who had evidence of previous COVID-19 infection (*P*= .045). Median titers were 104,354 (range 141-601,185), 113,608 (25-737,310), and 46,901 (25-694,528) for participants who were healthy, IMID not on methotrexate, and IMID on methotrexate, respectively. Similarly, in the Erlangen validation cohort, 179 of 182 (98.3%) of healthy controls, 10 of 11 (90.9%) of patients with IMID receiving no methotrexate, and 10 of 20 (50.0%) receiving methotrexate achieved adequate immunogenicity (Figure 1B). Median ODs for this cohort were 9.4 (range 1.2-14), 7.8 (2.3-11.3), and 5.9 (0.95-13.5) for participants who were healthy, IMID not on methotrexate, and IMID on methotrexate, respectively. Furthermore, when looking at the two cohorts in conjunction (n = 290), 204 of 208 (98.1%) of healthy controls, 34 of 37 (91.9%) of patients with IMID receiving no methotrexate, and 28 of 45 (62.2%) receiving methotrexate achieved adequate immunogenicity (*P*= < 0.001) (Figure S1).

**Figure 1.**
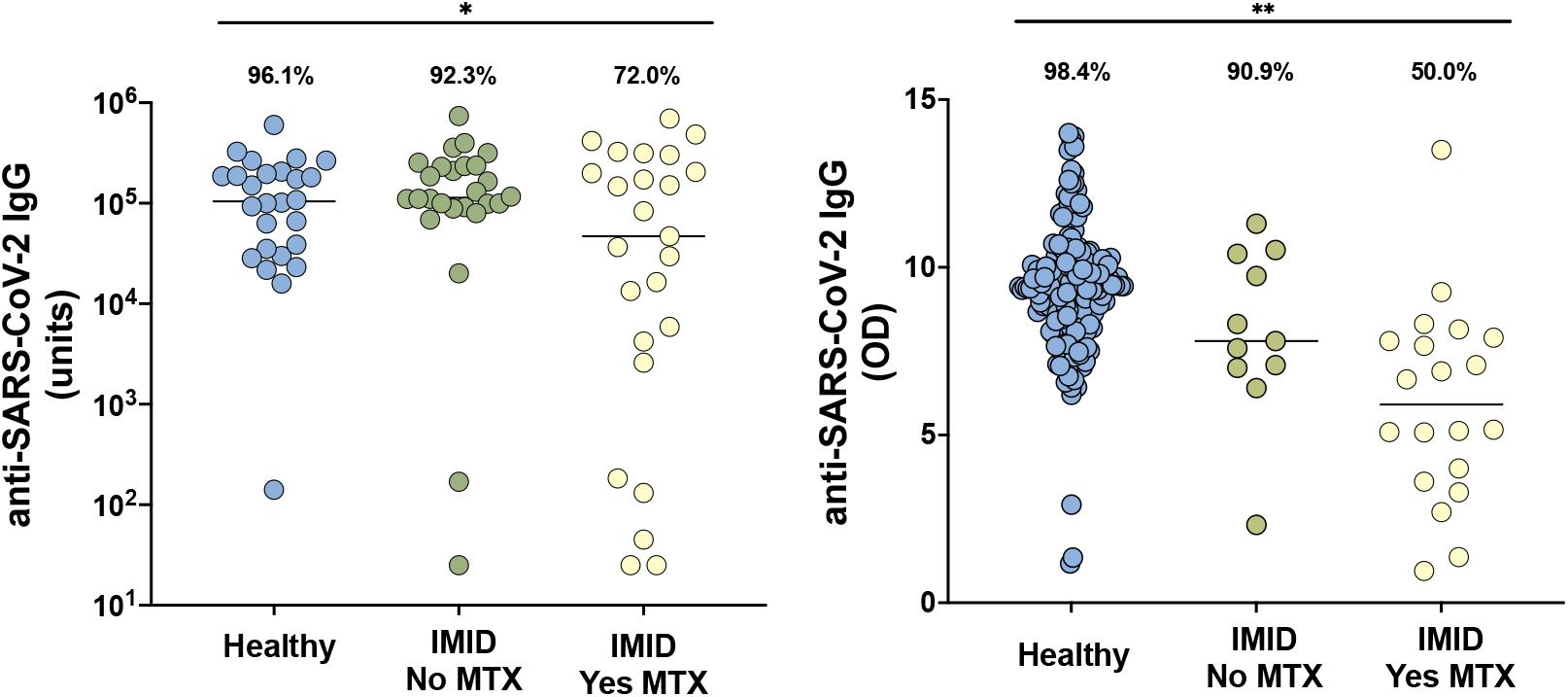
Anti-SARS-CoV-2 IgG levels in cohorts from New York City (A) and Erlangen (B) in healthy participants without immune-mediated inflammatory diseases (IMID; blue), IMID patients not receiving methotrexate (MTX; green), and IMID patients treated with MTX (yellow). Solid lines represent mean titer of each group. For the New York City cohort (A), adequate response is defined as greater than 5000 units and for the Erlangen cohort (B), adequate response is defined as greater than 5.7 (OD450nm), two standard deviations of the mean of controls. Percentages and group comparisons using chi squared test of independence reflect proportion of those achieving an adequate response within each group. * indicates p value less than .05. **indicated p value less than .001.

Because of the imbalance in age between groups, we further analyzed immunogenicity based on a cut off age of 55. In both age groups, the response rate for those on methotrexate remained significantly lower (*P*=< 0.001) (Figure S2). As an added sensitivity analysis, we used a stricter definition of inadequate antibody response (i.e., less than 1000 units for New York cohort and less than 5 OD for the Erlagen cohort). With the use of these more conservative cut off levels, patients with IMID on background methotrexate continued to show significantly decreased antibody response (*P*< .001) (Figure S3).

### Lack of CD8+ T-cell activation in IMID patients on methotrexate following mRNA COVID-19 vaccine

In the New York Cohort, 20 healthy controls, 24 patients with IMID not receiving methotrexate, and 18 patients with IMID who were receiving methotrexate underwent immune cell phenotyping before and after vaccination. The proportions of spike-specific B cells, circulating T follicular helper (cTfh; CD4+ICOS+CD38+ subset) cells, activated CD4+ T cells, and HLA-DR+ CD8+ T cells increased significantly in all groups after immunization (Figures 2A-D). Activated CD8+ T cells, defined as CD8+ T cells expressing Ki67 and CD38, and the granzyme B-producing (GZMB) subset of these activated CD8+ T cells, were induced in healthy adults and participants with IMID not on methotrexate, but not induced in patients receiving methotrexate (Figures 2E-F).

**Figure 2.**
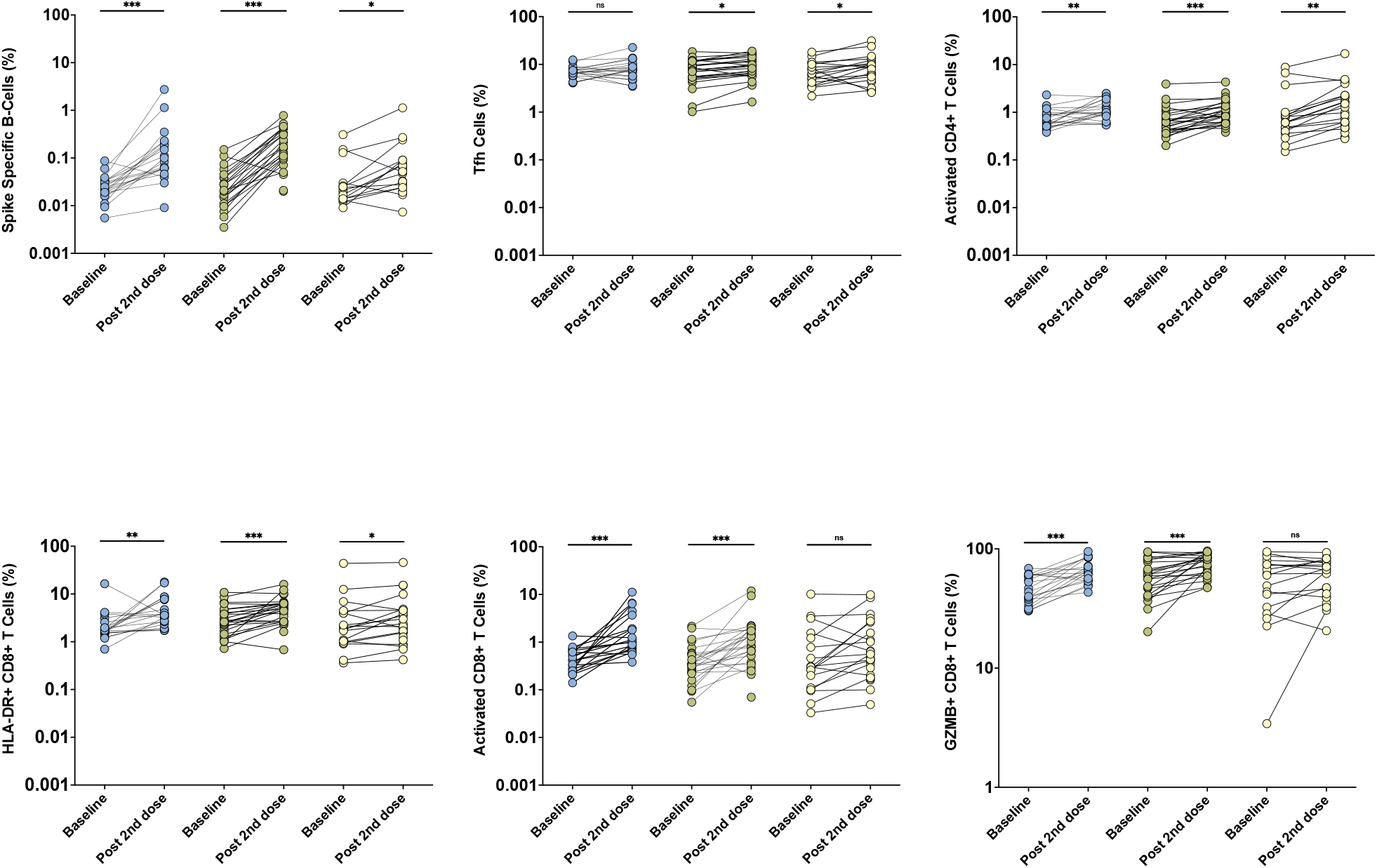
Immune cell populations from the New York City cohort by high-spectral flow in healthy controls (blue, n=20), patients with immune mediate inflammatory disease (IMID) not on methotrexate (MTX; green, n= 24), and patients with IMID on MTX (yellow, n=18), at baseline and post 2^nd^ dose of BNT162b2 mRNA vaccine. Pre- and post-vaccination comparisons were performed using Wilcoxon Rank Sum tests. Y-axes presented as a logarithmic scale. ns indicated no statistical significance * indicates p value less than .05 ** indicates p value less than .01 *** indicates p value less than .0001

## DISCUSSION

In two geographically independent cohorts of IMID patients, we found that methotrexate, a widely used immunomodulator for the treatment of several IMIDs, adversely affected humoral and cellular immunogenicity to COVID-19 mRNA vaccines.

For humoral immunity, the BNT162b2 mRNA vaccines did not induce adequately elevated SARS-CoV-2-spike-specific-IgG antibody titers in up to a third of the patients on methotrexate, as compared to IMID patients on other DMARDs, whom demonstrated a response as robust as that of healthy controls. This finding was analogous to the previously described effects of methotrexate on influenza vaccine immunogenicity.(5, 12-14) While a recent report has shown no differences in immunogenicity for patients with IMID, none of the included participants were on methotrexate.(15) A second study in patients with self-reported rheumatic and musculoskeletal diseases recruited via social media showed that 10 out of 13 participants on background methotrexate had detectable antibody levels after only one dose of SARS-CoV-2 mRNA vaccine,(16) although this was both underpowered and using a semi-quantitative ELISA measuring antibodies against SARS-CoV-2 receptor-binding domain. Therefore, the findings from our work looking at antibody responses in IMID patients after full vaccination regimen are of potentially high clinical relevance, since it was recently shown that a temporary discontinuation of methotrexate for two weeks significantly improved influenza vaccine immunogenicity in patients with rheumatoid arthritis.(2)

Importantly, the use of high-dimensional spectral flow cytometry allowed for the interrogation of specific cellular immune responses before and after immunization. Spike-specific B cells, activated CD4+ T cells and cTfh cells were induced similarly in all groups after mRNA vaccination. In contrast, activated CD8+ T cell responses were notably attenuated in the methotrexate cohort. Moreover, the poor induction of activated CD8+ T cells expressing granzyme B may indicate reduced cytotoxic functionality of these cells. Indeed, CD8+ T cell responses were identified to be a correlate of protection in non-human primate studies of SARS-CoV-2 infection.(17) Thus, reduced induction of cytotoxic CD8+ T cell responses, combined with inconsistent induction of antibody responses, may further impair effectiveness of COVID-19 vaccines and render IMID patients on methotrexate more at risk of inadequate vaccine response. However, this finding requires a cautious interpretation as it is quite possible that the use of methotrexate may delay (rather than prevent) adequate cellular mediated immunity against SARS-CoV-2. While spike-specific T-cell immunity has been detected as early as 10 days following one dose of mRNA COVID-19 vaccines in healthy individuals,(18) mRNA-1273-specific CD4+ and CD8+ T cell responses were most robustly elicited two weeks after the second dose.(19) Therefore, more detailed and comprehensive studies that include longer term characterization of the dynamics of cellular responses to these vaccines will be required to understand the clinical implications of these findings.

Although our analysis was limited in sample size, followed participants with biosampling for a relatively short period of time without standardized disease activity status metrics, and was restricted to one type of mRNA immunization, our findings were validated in an independent cohort and revealed that methotrexate, which is widely used for many indications, adversely affected the humoral and cellular immunogenicity to COVID-19 mRNA vaccination.

Furthermore, because of the inclusion of patients with prior COVID-19 infection, it is possible that results could be biased in favor of those not on methotrexate. However, when excluding all patients with prior infection, the results remained similar. We also acknowledge that there may have been participants with asymptomatic COVID-19 infection that we have not captured.

While immunosenescence may reduce the level of antibody responses to immunizations,(20) recent studies on COVID-19 mRNA vaccines have not shown differences in clinical outcomes for the older population.(6) In our study, IMID patients on methotrexate were generally older which may potentially explain some differences in immunogenicity. However, even when looking at participants younger than 55, decreased rates of humoral response were still significant. Further validation in even larger cohorts that address efficacy will be required to understand the interaction between age and methotrexate in the context of COVID-19 vaccination.

Importantly, it is not yet clear what level of immunogenicity is representative of vaccine efficacy (and this includes the arbitrary cut offs chosen for our measurements). We recognize that the definition of adequate cellular and humoral immune response may need to be refined in the future when correlation with efficacy becomes available. However, even after applying more conservative cut offs, the hampering effects of methotrexate on immunogenicity are still evident.

Taken together, our results suggest that the optimal protection of patients with IMID against COVID-19 will require further studies to determine whether additional doses of vaccine, dose modification of methotrexate, or even temporary discontinuation of this drug can boost immune response as has been demonstrated for other viral vaccines in this patient population.(7)

## Data Availability

All data relevant to the study are included in the article or uploaded as supplementary information. Further de-identified data can be made available upon request.

## Contributorship

R.H.H. and J.U.S designed the New York study, designed the data collection tools, analyzed and cleaned the data, and drafted and revised the paper. R.S.H, M.S., and M.J.M designed the New York study, designed, and performed the cellular analysis, and revised the paper. S.B. A. designed the New York Study and revised the paper. R.B.B. designed the New York study, acquired data, and revised the draft. D.S., R.A., K.T, M.N. and G.S. designed the Erlangen study, designed the data collection tools, analyzed and cleaned the data, and revised the paper. S.A. aided in original design, statistical analysis, and revised the paper. M.T, S.B.K., J.R.A., and A.R.C analyzed data and revised the draft. R.C., P.R., G. S., N.A, P.R., P.I., J.S., B.G., and S.R. helped accrue data and revised the draft.

## Acknowledgements

We would like to thank our patients and their families for participating in this study. We are grateful to Luz Alvarado, Rhina Medina, Parvathi Girija, Jyoti Patel, and Zakwan Uddin for coordinating and for data entry efforts. We would also like to thank Rebecca Cohen for regulatory support.

## Ethical Approval

New York University IRB 20-01078, University Clinic of Erlangen IRB 157_20B

## REFERENCES

1. Caldera F, Hillman L, Saha S, et al. Immunogenicity of High Dose Influenza Vaccine for Patients with Inflammatory Bowel Disease on Anti-TNF Monotherapy: A Randomized Clinical Trial. Inflammatory Bowel Diseases. 2019;26(4):593–602.

2. Park JK, Lee MA, Lee EY, et al. Effect of methotrexate discontinuation on efficacy of seasonal influenza vaccination in patients with rheumatoid arthritis: a randomised clinical trial. Annals of the Rheumatic Diseases. 2017;76(9):1559–65.

3. Kobie JJ, Zheng B, Bryk P, et al. Decreased influenza-specific B cell responses in rheumatoid arthritis patients treated with anti-tumor necrosis factor. Arthritis Research & Therapy. 2011;13(6):R209.

4. Hagihara Y, Ohfuji S, Watanabe K, et al. Infliximab and/or immunomodulators inhibit immune responses to trivalent influenza vaccination in adults with inflammatory bowel disease. Journal of Crohn’s and Colitis. 2014;8(3):223–33.

5. França ILA, Ribeiro ACM, Aikawa NE, et al. TNF blockers show distinct patterns of immune response to the pandemic influenza A H1N1 vaccine in inflammatory arthritis patients. Rheumatology. 2012;51(11):2091–8.

6. Polack FP, Thomas SJ, Kitchin N, et al. Safety and Efficacy of the BNT162b2 mRNA Covid-19 Vaccine. New England Journal of Medicine. 2020;383(27):2603–15.

7. Park JK, Lee YJ, Shin K, et al. Impact of temporary methotrexate discontinuation for 2 weeks on immunogenicity of seasonal influenza vaccination in patients with rheumatoid arthritis: a randomised clinical trial. Annals of the Rheumatic Diseases. 2018;77(6):898–904.

8. Haberman R, Axelrad J, Chen A, et al. Covid-19 in Immune-Mediated Inflammatory Diseases - Case Series from New York. N Engl J Med. 2020;383(1):85–8.

9. Amanat F, Stadlbauer D, Strohmeier S, et al. A serological assay to detect SARS-CoV-2 seroconversion in humans. Nature Medicine. 2020;26(7):1033–6.

10. Samanovic MI, Cornelius AR, Wilson JP, et al. Poor antigen-specific responses to the second BNT162b2 mRNA vaccine dose in SARS-CoV-2-experienced individuals. medRxiv. 2021:2021.02.07.21251311.

11. Simon D, Tascilar K, Krönke G, et al. Patients with immune-mediated inflammatory diseases receiving cytokine inhibitors have low prevalence of SARS-CoV-2 seroconversion. Nature Communications. 2020;11(1):3774.

12. Ribeiro ACM, Guedes LKN, Moraes JCB, et al. Reduced seroprotection after pandemic H1N1 influenza adjuvant-free vaccination in patients with rheumatoid arthritis: implications for clinical practice. Annals of the Rheumatic Diseases. 2011;70(12):2144–7.

13. Adler S, Krivine A, Weix J, et al. Protective effect of A/H1N1 vaccination in immune-mediated disease—a prospectively controlled vaccination study. Rheumatology. 2011;51(4):695–700.

14. Hua C, Barnetche T, Combe B, et al. Effect of Methotrexate, Anti–Tumor Necrosis Factor α, and Rituximab on the Immune Response to Influenza and Pneumococcal Vaccines in Patients With Rheumatoid Arthritis: A Systematic Review and Meta-Analysis. Arthritis Care & Research. 2014;66(7):1016–26.

15. Geisen UM, Berner DK, Tran F, et al. Immunogenicity and safety of anti-SARS-CoV-2 mRNA vaccines in patients with chronic inflammatory conditions and immunosuppressive therapy in a monocentric cohort. Annals of the Rheumatic Diseases. 2021:annrheumdis-2021- 220272.

16. Boyarsky BJ, Ruddy JA, Connolly CM, et al. Antibody response to a single dose of SARS-CoV-2 mRNA vaccine in patients with rheumatic and musculoskeletal diseases. Ann Rheum Dis. 2021.

17. McMahan K, Yu J, Mercado NB, et al. Correlates of protection against SARS-CoV-2 in rhesus macaques. Nature. 2021;590(7847):630–4.

18. Kalimuddin S, Tham CY, Qui M, et al. Early T cell and binding antibody responses are associated with Covid-19 RNA vaccine efficacy onset. Med (N Y). 2021.

19. Jackson LA, Anderson EJ, Rouphael NG, et al. An mRNA Vaccine against SARS-CoV-2 Preliminary Report. N Engl J Med. 2020;383(20):1920–31.

20. Walsh EE, Frenck RW, Falsey AR, et al. Safety and Immunogenicity of Two RNA-Based Covid-19 Vaccine Candidates. New England Journal of Medicine. 2020;383(25):2439–50.

