## Supplemental Appendix for "Methotrexate Hampers Immunogenicity to BNT162b2 mRNA COVID-19 Vaccine in Immune-Mediated Inflammatory Disease"

### Supplementary Appendix

### TABLE OF CONTENTS

Table S1. Characteristics of cohort from Erlangen, Germany Page 2
Figure S1. Page 3
Figure S2. Page 4
Figure S3. Page 5
Supplementary Methods
Anti-SARS-CoV-2 IgG antibody titers Page 6
Immune cell phenotyping by high-dimensional spectral flow cytometry Page 6
Table S2. Antibodies used for high-dimensional spectral flow cytometry Page 7
Figure S3 Page 8

**Supplementary Table 1**. Characteristics of cohort from Erlangen, Germany.

| Characteristic | Controls (n = 182) | IMID No MTX⁺ (n =11) | IMID Yes MTX⁺ (n = 20) | p-value |
|---|---|---|---|---|
| **Age- mean (range, SD)** | 40.8 [21-65, 12.0] | 45.0 [26-80, 15.5] | 54.5 [24-87, 19.2] | 0.008 |
| **Female- n (%)** | 104 (57.1) | 7 (58.3) | 15 (75.0) | 0.290 |
| **Race- n (%)** | | | | |
| White | 178 (97.8) | 11 (100.0) | 19 (95.0) | 0.680 |
| Black | 0 (0.0) | 0 (0.0) | 0 (0.0) | |
| Asian | 2 (1.1) | 0 (0.0) | 1 (5.0) | |
| Other | 1 (0.6) | 0 (0.0) | 0 (0.0) | |
| **Hispanic ethnicity- n (%)** | 1 (0.6) | 0 (0.0) | 0 (0.0) | 0.918 |
| **Long term medication use- n (%)** | | | | |
| Methotrexate | -- | 0 (0.0) | 20 (100.0) | 1.00 |
| Tumor necrosis factor inhibitors | -- | 11 (100.0) | 0 (0.0) | 1.00 |
| **Methotrexate dose- mean (SD)** | -- | -- | 14.7 (4.2) | |
| **COVID-19 infection prior to vaccination- n(%)** | 0 (0.0) | 0 (0.0) | 0 (0.0) | 1.00 |
| **Days from 1st vaccine mean (range, SD)** | 47.7 (11-78,12.1) | 42.1 (23-74,18.0) | 44.0 (13-78,16.7) | 0.204 |
| **Number receiving 2nd vaccination dose- n (%)** | 179 (98.4) | 9 (81.8) | 18 (90.0) | 0.044 |
| **Adequate humoral immunogenicity#- n (%)** | 179 (98.4) | 10 (90.9) | 10 (50.0) | <0.001 |
| **OD anti-SARS-CoV-2 IgG** | | | | |
| Median (range) | 9.4 (1.2-14) | 7.8 (2.3-11.3) | 5.9 (0.95-13.5) | <0.001 |

⁺IMID denotes immune-mediated inflammatory disease, MTX methotrexate, OD optical density.
*Giant cell arteritis and polymyalgia rheumatica.
#Adequate response defined as within 2 standard deviations (SD) of the mean OD450nm of controls (OD 450nm >5.7units).

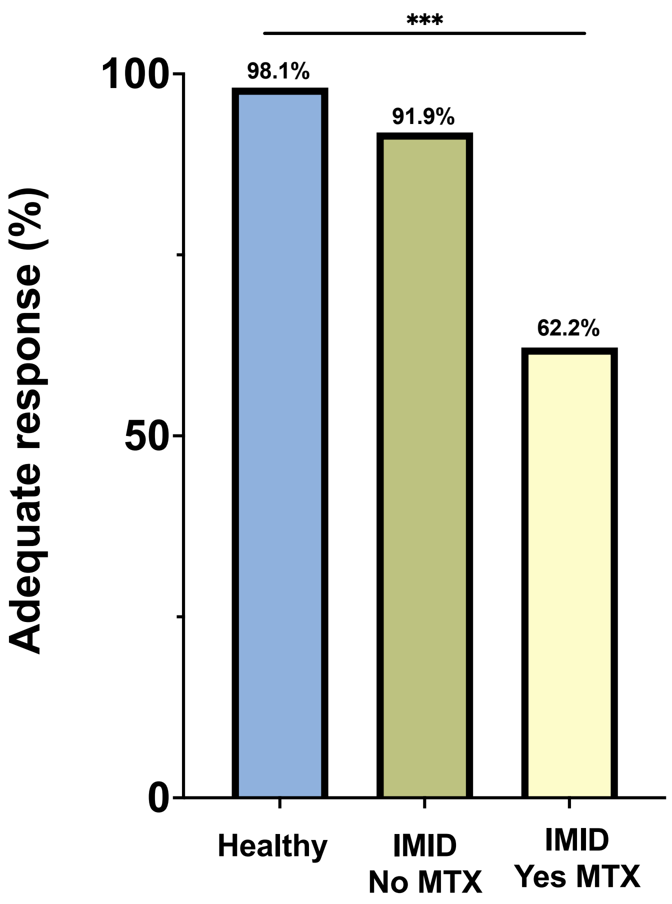


**Supplementary Figure 1.** Percentage of patients achieving adequate response to COVID-19 vaccination from New York and Erlangen cohorts combined (n=290). Percentages and group comparisons using chi squared test of independence reflect proportion of those achieving an adequate response within each group.
***indicated p value less than .001.

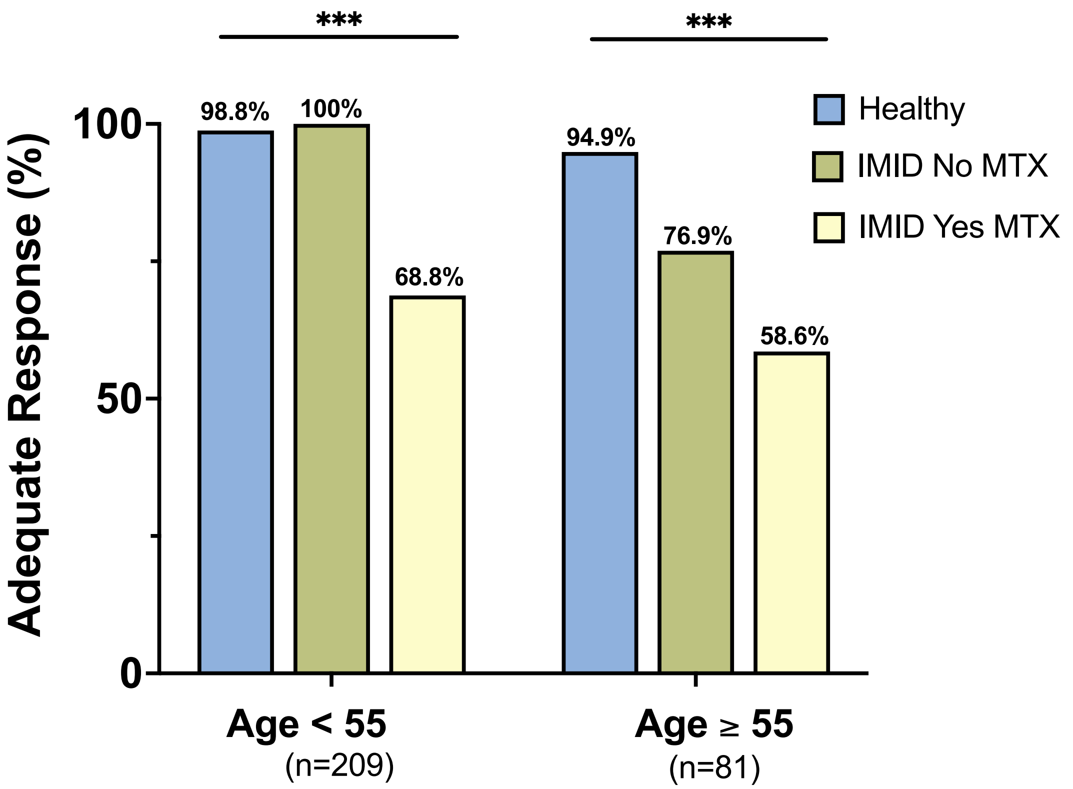


**Supplementary Figure 2.** Percentage of total patients achieving adequate response to COVID-19 vaccination by age. Percentages and group comparisons using chi squared test of independence reflect proportion of those achieving an adequate response within each group.
***indicated p value less than .001.

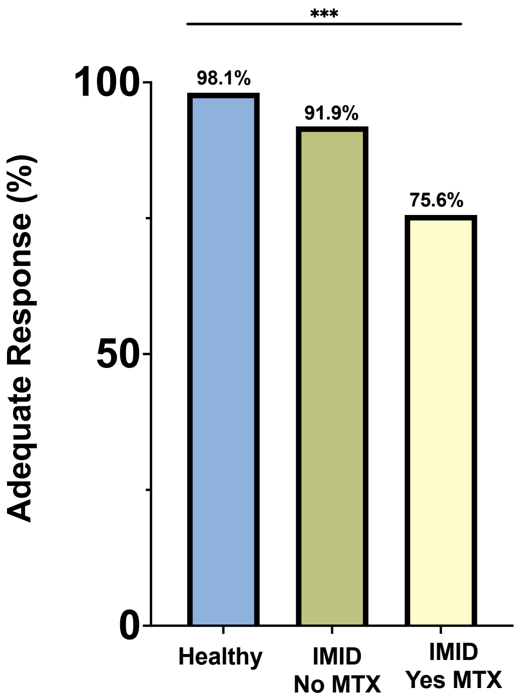


**Supplementary Figure 3.** Percentage of total patients achieving a stricter definition of adequate response to COVID-19 vaccination. For the New York City cohort, adequate response is defined as greater than 1000 units and for the Erlangen cohort, adequate response is defined as greater than 5.0 (OD450nm). Percentages and group comparisons using chi squared test of independence reflect proportion of those achieving an adequate response within each group.
***indicated p value less than .001.

### Supplementary Methods

#### Anti-SARS-CoV-2 IgG antibody titers

Direct ELISA was used to quantify antibody titers in the New York cohort participants serum. Ninety-six well plates were coated with 1 µg/ml S1 protein (100 µl/well) diluted in PBS and were then incubated overnight at 4°C (Sino Biological Inc., 40591-V08H). Plates were washed four times with 250 µl of PBS containing 0.05% Tween 20 (PBS-T) and blocked with 200 µl PBS-T containing 4% non-fat milk and 5% whey, as blocking buffer at RT for 1 hour. Sera were heated at 56°C for 1 hour prior to use. Samples were diluted to a starting concentration of 1:50 (S1) were first added to the plates and then serially diluted 1:3 in blocking solution. The final volume in all wells after dilution was 100 µl. After a 2-hour incubation period at RT, plates were washed four times with PBS-T. Horseradish-peroxidase conjugated goat-anti human IgG (Southern BioTech, 2040-05) were diluted in blocking buffer (1:2000) and 100 µl was added to each well. Plates were incubated for 1 hour at RT and washed four times with PBS-T. After developing for 5 min with TMB Peroxidase Substrate 3,3′,5,5′-Tetramethylbenzidine (Thermo Scientific), the reaction was stopped with 1M sulfuric acid or 1N hydrochloric acid. The optical density was determined by measuring the absorbance at 450 nm on a Synergy 4 (BioTek) plate reader. In order to summarize data collected on individuals, the area under the response curve was calculated for each participant and end point titers were normalized using replicates of pooled positive control sera on each plate to reduce variability between plates. IgG antibodies against the S1 domain of the spike protein of SARS-CoV-2 were tested in the Erlangen participants using the commercial enzyme–linked immunosorbent assay from Euroimmun (Lübeck, Germany) on the EUROIMMUN Analyzer I platform and according to the manufacturers protocol. Optical density was determined at 450 nm with reference wavelength at 630 nm.

#### Immune cell phenotyping by high-dimensional spectral flow cytometry

Peripheral blood was collected in sodium heparin collection tubes and PBMC were isolated using the SepMate system in accordance with manufacturer's instructions. PBMC were cryopreserved in fetal calf serum supplemented with 10% DMSO. Cryopreserved cells were thawed in batches for immunophenotyping studies. Then, 2 to 5 million freshly isolated PBMC were resuspended in HBSS supplemented with 1% fetal calf serum (Fisher) and 0.02% sodium azide (Sigma). Cells underwent Fc-blockade with Human TruStain FcX (Biolegend) and NovaBlock (Phitonex) for 10 minutes at room temperature, followed by surface staining antibody cocktail at room temperature for 20 minutes in the dark. Cells were permeabilized with the eBioscience Intracellular Fixation and Permeabilization kit (Fisher) for 20 minutes at room temperature in the dark, followed by intracellular staining with an antibody cocktail for 1 hour at room temperature in the dark. All samples were then resuspended in 1% paraformaldehyde and acquired within three days of staining on a 5-laser Aurora cytometer (Cytek Biosciences). Antibodies, clones, and catalog numbers are available in Table S3. Initial data quality control was performed using FlowJo.

**Supplementary Table 2.** Antibodies used for high-dimensional spectral flow cytometry

| Target | Fluorochrome | Clone | Manufacturer | Catalog # |
|---|---|---|---|---|
| **Live/Dead Blue** | - | - | Invitrogen | L23105 |
| **CD3** | APC/Fire 810 | SK7 | Biolegend | 344857 |
| **CD4** | SparkBlue 550 | SK3 | Biolegend | 344656 |
| **CD8** | PE-Fire 640 | SK1 | Biolegend | 344761 |
| **CD11c** | PerCP | Bu15 | Biolegend | 337234 |
| **CD14** | BUV805 | M5E2 | BD | 612902 |
| **CD16** | BV480 | 3G8 | BD | 566171 |
| **CD19** | BUV496 | SJ25C1 | BD | 612939 |
| **CD20** | APC | 2H7 | Biolegend | 302309 |
| **CD21** | PE-Cy5 | B-ly4 | BD | 551064 |
| **CD23** | BUV615 | M-L233 | BD | 751104 |
| **CD25** | BUV563 | 2A3 | BD | 612919 |
| **CD27** | SB702 | O323 | Invitrogen | 67-0279-42 |
| **CD38** | Qdot655 | HIT2 | Invitrogen | Q22150 |
| **CD40** | BV510 | 5C3 | Biolegend | 334330 |
| **CD45RA** | Spark NIR 685 | HI100 | Biolegend | 304168 |
| **CD56** | BV570 | 5.1H11 | Biolegend | 362539 |
| **CD71** | SB780 | OKT9 | Invitrogen | 78-0719-42 |
| **CD123** | BV650 | 7G3 | BD | 563405 |
| **Recombinant Spike protein** | PE | | Biolegend | 793804 |
| **CD138** | PacBlue | MI15 | Biolegend | 356531 |
| **CD150 (CTLA4)** | BV421 | BNI3 | Biolegend | 369606 |
| **CD183 (CXCR3)** | BV750 | 1C6 | BD | 746895 |
| **CD185 (CXCR5)** | BB515 | RF8B2 | BD | 564624 |
| **CD197 (CCR7)** | BV605 | G043H7 | Biolegend | 353224 |
| **CD278 (ICOS)** | APC-Fire750 | C398.4A | Biolegend | 313536 |
| **CD279 (PD-1)** | BB700 | EH12.1 | BD | 566460 |
| **HLA-DR** | BUV661 | G46-6 | BD | 612980 |
| **IgD** | BUV737 | IA6-2 | BD | 612798 |
| **Foxp3** | PE-Cy5.5 | PCH101 | Invitrogen | 35-4776-42 |
| **Tbet** | PE-Cy7 | 4B10 | Biolegend | 644823 |
| **Eomes** | PE-eF610 | WD1928 | Invitrogen | 61-4877-42 |
| **GzmB** | A700 | GB11 | BD | 561016 |
| **Ki67** | BUV395 | B56 | BD | 564071 |
| **IgG** | PerCP-Vio700 | IS11-3B2.2.3 | Miltenyi | 130-119-880 |

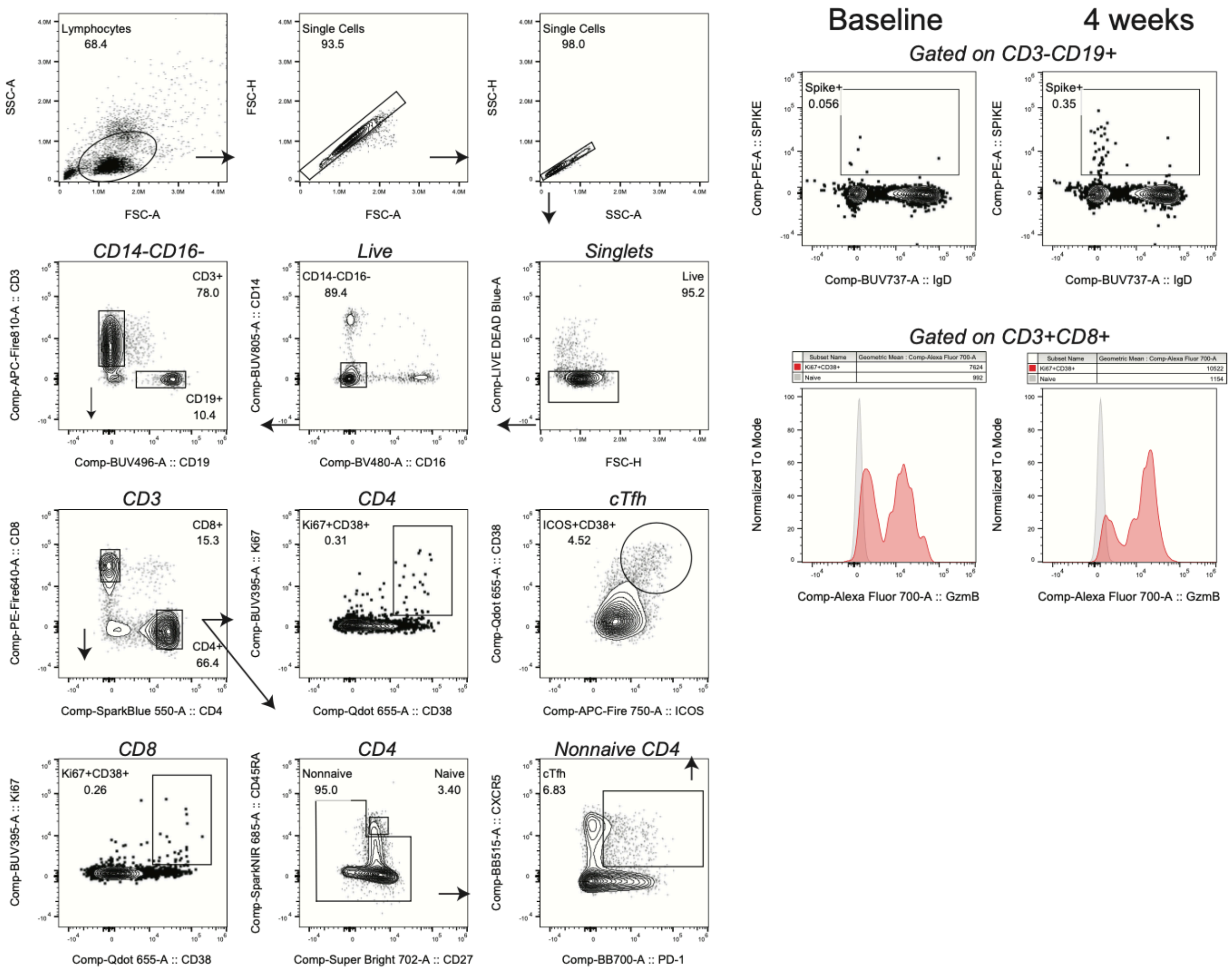


**Supplementary Figure 4.** Flow cytometry gating strategy. (Left panel) Gating strategy and example plots for the key immune populations. (Right panel) Plots showing baseline or at the 4-week time point for CD19 specific for Spike protein (top row) or for expression of Granzyme B in CD8 T cells
